# Early Carotid Revascularization Rates, Procedural Distribution, and Hospital Density

**DOI:** 10.1101/2022.04.26.22274347

**Authors:** Andrew Russeau, Deeksha Bidare, Kyle E. Walker, Panos Kougias, Joseph L. Mills, Neal R. Barshes

## Abstract

Early carotid revascularization (i.e. during the index hospitalization) may help reduce the risk of additional neurologic events without excess perioperative morbidity. We evaluated the relationship between rates of early carotid endarterectomy (CEA) or stenting (CAS) and hospital density/distribution within metropolitan areas of Texas. Patients with extracranial carotid artery stenosis and either stroke, transient ischemic attacks, or amaurosis were identified among all patients admitted from 2009 to 2013 to non-federal Texas hospitals within all 24 Texas metropolitan statistical areas (MSAs). Early CEA/CAS was defined as occurring during the index hospitalization. A Gini coefficient with bias correction factor was calculated to quantify the distribution of carotid procedures within an MSA. In total, 3,330 (15.4%) of the 21,665 metropolitan patients admitted to Texas hospitals with symptomatic carotid stenosis received early CEA/CAS. Only 263 (44%) of the 600 total hospitals where patients were admitted performed early CEA/CAS. An increasing proportion receiving early carotid CEA/CAS was inversely correlated with the procedural Gini coefficient (p=0.002) and directly correlated with the number of hospitals per 100K population (p=0.01). These two factors accounted for 51% of the variability among MSAs. Early CEA/CAS rates did not correlate with hospital volume or with level 1or 2 stroke centers within an MSA. Increasing the number of hospitals performing early carotid revascularization procedures (i.e. avoiding regionalization/concentration) may help increase the number of patients receiving early carotid revascularization for symptomatic carotid artery disease.

## Introduction

Delaying carotid endarterectomy (CEA) for 4-6 weeks after stroke or transient ischemic attack (TIA) had common practice at the time of the North American Symptomatic Carotid Endarterectomy Trial and the European Carotid Surgery Trial. Subsequent post-hoc analyses of data from these trials, however, suggested that the absolute risk reduction conferred by CEA decreases beyond the first two weeks following stroke or TIA.^1^ Incorporating findings from these analyses and many other data sources [including those summarized by Naylor]^2^ recommendations for the optimal timing of CEA/CAS have since changed to 2-14 days after the index neurological event.^3–5^

Access to hospitals offering early CEA/CAS, however, may be limited or uneven in many geographic areas of the United States. Analyses of persons in Texas with peripheral artery disease and limb threatening foot complications such as ulcers or gangrene have suggested that place of residence and hospital travel distance may influence -- and perhaps even constrain -- hospital choice and, as a result, access to revascularization to avoid leg amputation.^6^ With these considerations in mind, we sought to evaluate the factors affecting the rate of intervention in symptomatic carotid stenosis focusing on hospital density.

## Methods

We obtained data on all non-federal Texas hospital admissions between 2009 and 2013 from the Texas Department of State Health Services. No institutional review board approval was requested, as this is de-identified data available to the public. The study population of interest was adults living in metropolitan areas of Texas who were hospitalized for acute cerebral events due to lesions in the extracranial (cervical) portion of the carotid artery. The study sample was obtained by identifying patients age 45 or higher with diagnosis codes that identified both (1) carotid artery stenosis; and (2) stroke, transient ischemic attack, or amaurosis fugax using the ICD-9 diagnosis codes listed in Appendix 1. The ICD-9 procedure codes (Appendix 1) were then used to identify how many of these patients underwent early CEA/CAS, defined here as being performed during the index hospitalization. Patients who lived outside of metropolitan areas in Texas were excluded.

The patient-level data was then collated at the MSA level and combined with other data also aggregated at the MSA level, including: (1) population and demographics (from the US Census Bureau); (2) the number and per capita density of level 1 (comprehensive) and level 2 (primary) stroke centers as designated by The Joint Commission (Texas-specific list from the Texas Department of State Health Services); (3) distance travelled to receive treatment; and (4) the procedural Gini coefficient.

For each symptomatic carotid patient, latitude and longitude coordinates their zip code of residence centroid and the coordinates of the hospital at which they were treated were used with the Haversine formula (signed decimal degrees without compass direction^6^) to estimate distances travelled. The aggregated median distance traveled and proportion of patients who traveled more than 50 miles were calculated for each of the 24 Texas MSAs.

The Gini index or coefficient is most frequently used as a measure of income distribution.^7^ This has been applied to medicine to describe biodiversity, disease distribution, and regionalization of primary care.^8^ A Gini coefficient of 0 signifies perfect equality with all measured data points being equally distributed. Maximum inequality reflecting complete concentration is represented by a coefficient of 1. We calculated a procedural Gini coefficient for each MSA to quantify the degree of distribution vs concentration of carotid procedures within an MSA (Appendix 2).

We utilized this to determine the degree of regionalization of carotid intervention within each MSA, with a Gini coefficient of 0 signifying all hospitals in the MSA performed an equal number of carotid procedures while a coefficient of 1 means all procedures were performed at a single hospital. Additionally, the Herfindahl-Hirschman index was calculated as an additional measure of competition and distribution (Appendix 2).

Finally, the number and location of patients with symptomatic carotid disease was mapped relative to the location and procedural volume of hospitals. These maps were then assessed qualitatively to identify potential geospatial gaps or imbalances in CEA/CAS.

Intercooled Stata SE version 8.2, RStudio version 1.2.5033, Microsoft Excel version 16, and Google Sheets were used for statistical analyses. Mapping was done using the R packages *tigris* and *crsuggest* written by one of the authors (K.E.W.). Distance calculations were obtained using the *ggmap* package and the Google Geocoding API. The GINI coefficient was calculated using a Google Apps Script modified from the GINISS script by Dr. Paul Brewer.^9^ Descriptive statistics are presented as medians and 25-75% interquartile ranges (IQRs) unless otherwise stated. A p-value of <0.05 was considered statistically significant.

## Results

During the 5 years of the study period, a total of 27,132 patients 45 years of age or more presented to Texas hospitals with symptomatic carotid stenosis. In total, 21,665 (79.9%) resided in metropolitan areas of Texas and were included in the study. The median length of stay for these patients was 5 days (25-75% IQR of 3-8 days), with 17,217 (79.5%) having a length of stay of three or more days. Of these, 3,330 (15.4%) patients received early CEA/CAS. Upon aggregation of the patient-level data into 24 Texas MSAs, the median number of patients per MSA was 381.5 (IQR of 262.5 – 502.5).

Selected characteristics of MSAs are shown in Table 1. Wide ranges in the proportions of populations within various MSAs categorized as black (median 7.0%, range 0.3 to 24.2%) and Hispanic (median 29.9%, range of 6.8% to 95.8%) demonstrate the great racial and ethnic diversity present in the state of Texas. The median incidence rate of symptomatic carotid events per 100K population per year was 23.1 (IQR of 18.8 – 36.1) during the study period. Three MSAs (Austin-Round Rock, Brownsville-Harlingen, and Beaumont-Port Arthur) were found to have much higher incidence rates of symptomatic carotid events (74.0, 91.0 and 123.9 per 100K population per year, respectively). The median proportion of patients receiving early CEA/CAS per MSA was 15.1%, with an IQR of 13.3% - 20.1% and a range of 3.9% to 24.6%.

**Table 1:** Characteristics related to the treatment of symptomatic carotid disease for metropolitan statistical areas in Texas, 2009-2013.

| Metropolitan Statistical Area | Population in 2011 | categorized as black, % | categorized as Hispanic, % | hospitals per 10,000 persons | hospitalizations for symptomatic carotid disease, per 10,000 person-years | procedural Gini coefficient | median distance travelled to hospital, miles | received early carotid intervention, % |
| --- | --- | --- | --- | --- | --- | --- | --- | --- |
| Abilene | 166,185 | 7.0 | 21.8 | 9.0 | 25.3 | 0.92 | 8.2 | 20.0 |
| Amarillo | 256,494 | 5.8 | 25.8 | 2.7 | 21.4 | 0.89 | 4.9 | 13.8 |
| Austin-Round Rock | 1,769,045 | 7.0 | 31.8 | 2.8 | 74.0 | 0.86 | 8.4 | 15.2 |
| Beaumont-Port Arthur | 405,199 | 24.2 | 13.0 | 5.9 | 123.9 | 0.84 | 9.5 | 18.1 |
| Brownsville-Harlingen | 414,348 | 0.3 | 88.4 | 5.1 | 91.0 | 0.87 | 6.7 | 14.1 |
| College Station-Bryan | 232,801 | 11.6 | 23.1 | 6.0 | 16.5 | 0.95 | 5.9 | 14.6 |
| Corpus Christi | 434,648 | 3.1 | 58.3 | 4.8 | 23.6 | 0.92 | 8.5 | 16.6 |
| Dallas-Fort Worth | 6,571,761 | 14.7 | 28 | 1.8 | 19.3 | 0.8 | 6.6 | 15.0 |
| El Paso | 825,025 | 2.6 | 82.5 | 1.8 | 10.4 | 0.74 | 6.8 | 7.7 |
| Houston-Galveston | 6,057,425 | 16.8 | 36.1 | 1.5 | 17.3 | 0.81 | 8.0 | 17.9 |
| Killeen-Temple-Fort Hood | 414,000 | 18.5 | 20.8 | 4.8 | 19.6 | 0.97 | 10.7 | 13.3 |
| Laredo | 259,539 | 0.2 | 95.8 | 4.6 | 13.4 | 0.89 | 9.4 | 12.1 |
| Longview | 216,896 | 17.2 | 14.8 | 9.2 | 35.2 | 0.87 | 11.5 | 24.6 |
| Lubbock | 295,152 | 7.0 | 33.2 | 3.4 | 29.7 | 0.89 | 7.8 | 24.0 |
| McAllen-Edinburg | 798,286 | 0.4 | 90.8 | 2.5 | 11.8 | 0.92 | 6.8 | 14.9 |
| Midland-Odessa CSA* | 284,538 | 5.1 | 46.1 | 4.6 | 20.5 | 0.81 | 6.3 | 17.5 |
| San Angelo | 112,642 | 3.6 | 36.1 | 8.0 | 33.4 | 0.81 | 9.1 | 20.2 |
| San Antonio | 2,197,139 | 6.1 | 54.4 | 2.5 | 15.3 | 0.84 | 6.8 | 12.4 |
| Sherman-Denison | 121,800 | 5.8 | 11.8 | 13.1 | 47.5 | 0.75 | 4.8 | 13.1 |
| Texarkana | 93,275 | 24.2 | 6.8 | 6.4 | 38.8 | 0.83 | 7.4 | 3.9 |
| Tyler | 213,578 | 17.7 | 17.9 | 6.1 | 32.6 | 0.94 | 7.3 | 21.6 |
| Victoria | 94,693 | 5.9 | 43.8 | 8.4 | 22.2 | 0.84 | 1.2 | 23.8 |
| Waco | 256,237 | 15.2 | 24.1 | 4.7 | 22.6 | 0.89 | 7.9 | 8.3 |
| Wichita Falls | 151,299 | 8.9 | 15.7 | 5.9 | 40.5 | 0.97 | 8.6 | 20.3 |
| *median* | *272,039* | *7.0* | *29.9* | *4.8* | *23.6* | *0.87* | *7.6* | *15.1* |

\*CSA, combined statistical area

263 hospitals (44% of all Texas hospitals treating patients with symptomatic carotid events) performed early CEA/CAS with 201 hospitals (33%) performing at least 10 of these procedures. The total hospitals per MSA ranged from 6 to 118, with a median hospital density of 4.8 per 100K population (range 1.8 to 13.1 hospitals per 100K population). When limited to hospitals performing more than 10 early CEA/CAS procedures, the number per 100K ranged from 0.7 to 2.5. Each hospital had 1-730 (mean 35, median 2) patients admitted with symptomatic carotid disease. No significant association was found between hospital volume and the rate of early CEA/CAS. The rate of early CEA/CAS had no significant correlation with the per capita density of level 1 or level 2 stroke facilities as designated by The Joint Commission.

The Gini coefficients of early CEA-CAS ranged from 0.53 to 0.99 (median 0.82, IQR 0.76-0.87). The median distance travelled per MSA ranged from 1.23 to 11.47 (median 7.58, IQR 6.68 - 8.56). The proportion of patients who traveled greater than 50 miles ranged from 0.9% to 19.1% (median 2.9%, IQR 2.1% - 5.3%). No significant correlation was seen between proportion receiving early CEA/CAS and these measures of distance traveled by patients.

The final regression model demonstrated that higher proportions of patients receiving early carotid intervention were significantly associated with the number of hospitals per 100K population within a MSA (p=0.001) and with lower procedural Gini coefficients (i.e. a more even distribution of carotid interventions within a region rather than concentration at fewer hospitals; p=0.002; see Figure 1 and Table 2). The overall model R^2^ value was 0.50, suggesting that half of the inter-MSA variability in early CEA/CAS was explained by these two factors. No significant correlation was seen between the proportion undergoing early CEA/CAS within an MSA and any of the following characteristics: MSA demographics (including proportion of the population categorized as black or Hispanic); median distance travelled or proportion travelling >50 miles to the hospital at which they were treated; the incidence rate of symptomatic carotid events within the MSA; the ratio of symptomatic carotid events to hospitals; or the Herfindahl-Hirschman index.

**Table 2:** Factors independently and significantly associated with the proportion of persons hospitalized in Texas with symptomatic carotid disease who undergo early carotid intervention. Overall model R is 0.50.

| Factor | β-coefficient | 95% confidence interval | t value | p value |
| --- | --- | --- | --- | --- |
| procedural Gini coefficient | -0.464 | -0.733 to -0.194 | -3.6 | 0.002 |
| hospitals per 100,000 population | 0.011 | 0.003 to 0.019 | 2.8 | 0.010 |

**Figure 1:**
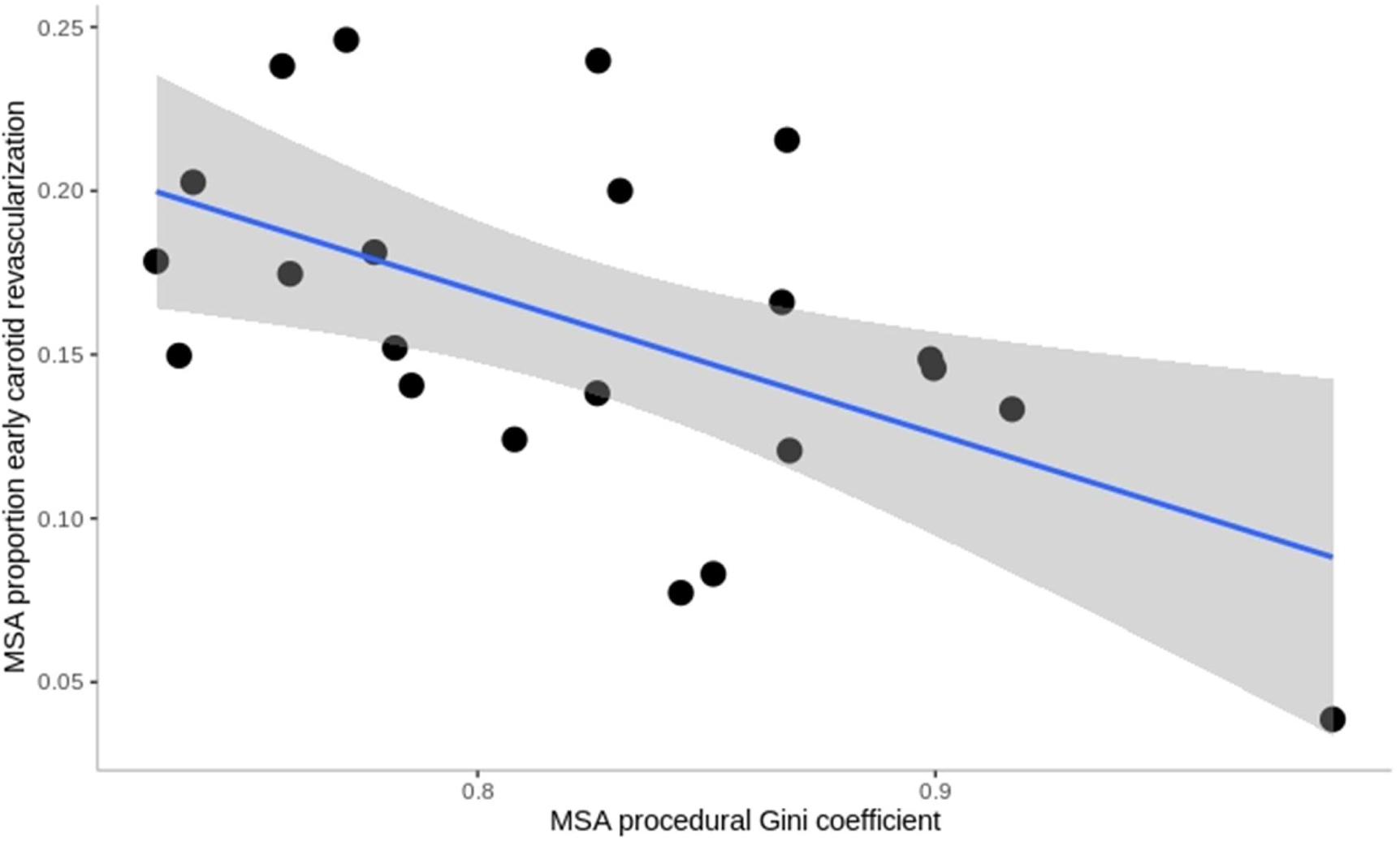
Scatterplot demonstrating the relationship between procedural Gini coefficient (horizontal axis) and rate of early carotid revascularization (vertical axis).

The location of symptomatic carotid stenosis patients relative to hospital location was analyzed in selected MSAs with specialized mapping (see Figures 2 and 3 for examples). Virtually zip codes in the highest quartile for the total number of symptomatic carotid stenosis patients had a hospital within the same zip code. Together with the results of the aforementioned MSA-level regression analysis and distance calculations, this suggested that location or distance may not influence the rate of early CEA/CAS as much as the relative distribution of procedures within an MSA.

**Figure 2:**
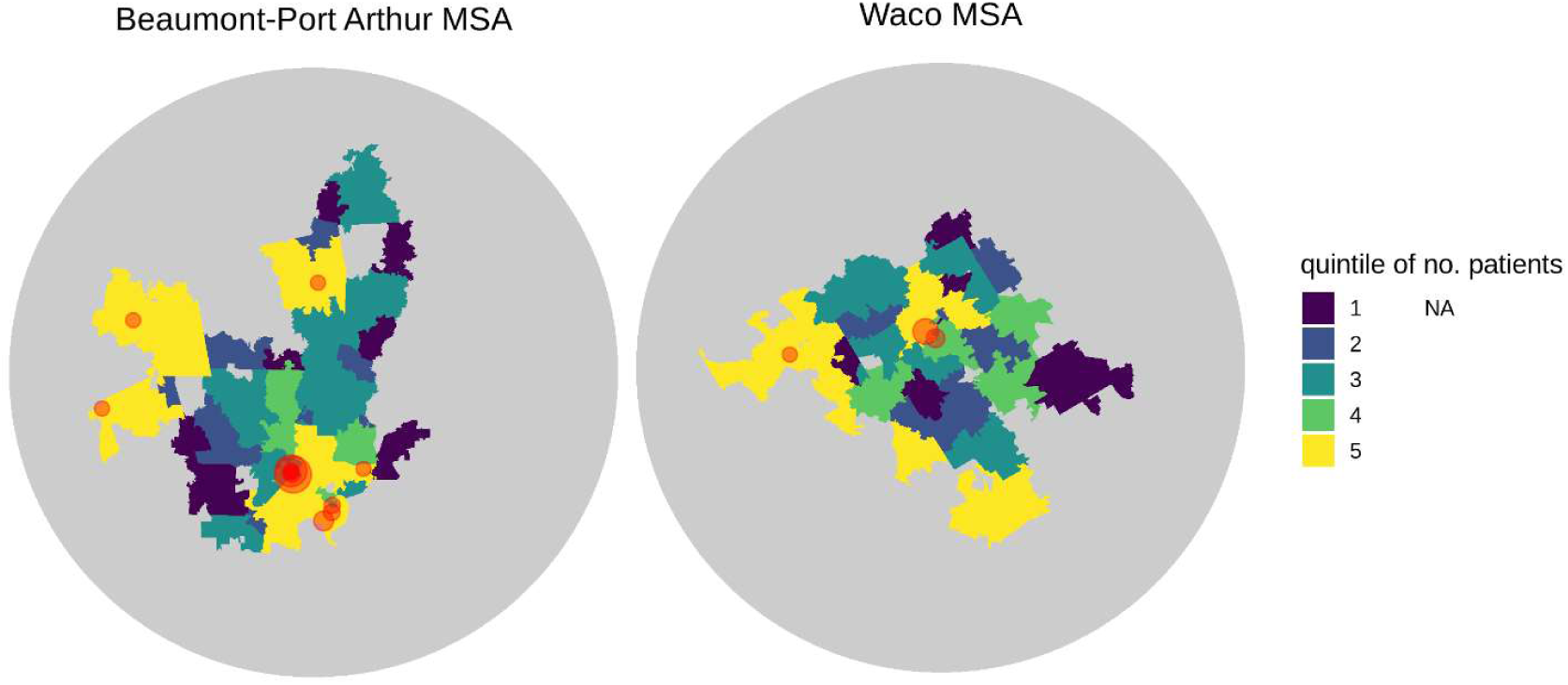
Map of the Beaumont-Port Arthur (left) and Waco (right) metropolitan statistical areas as examples of mid-sized cities. Color coding shows quintile of absolute number of patients with symptomatic carotid disease per zip code. Semi-transparent red circles demonstrate location of hospitals performing CEA/CAS, with circle size reflecting procedural volume. Compared to Beaumont-Port Arthur, the hospitals performing early carotid revascularization in Waco are fewer and more concentrated in location. There are two top-quartile zip codes in Waco with no hospitals performing early carotid revascularization, whereas in Beaumont-Port Arthur there are no top-quartile zip codes without such hospitals.

**Figure 3:**
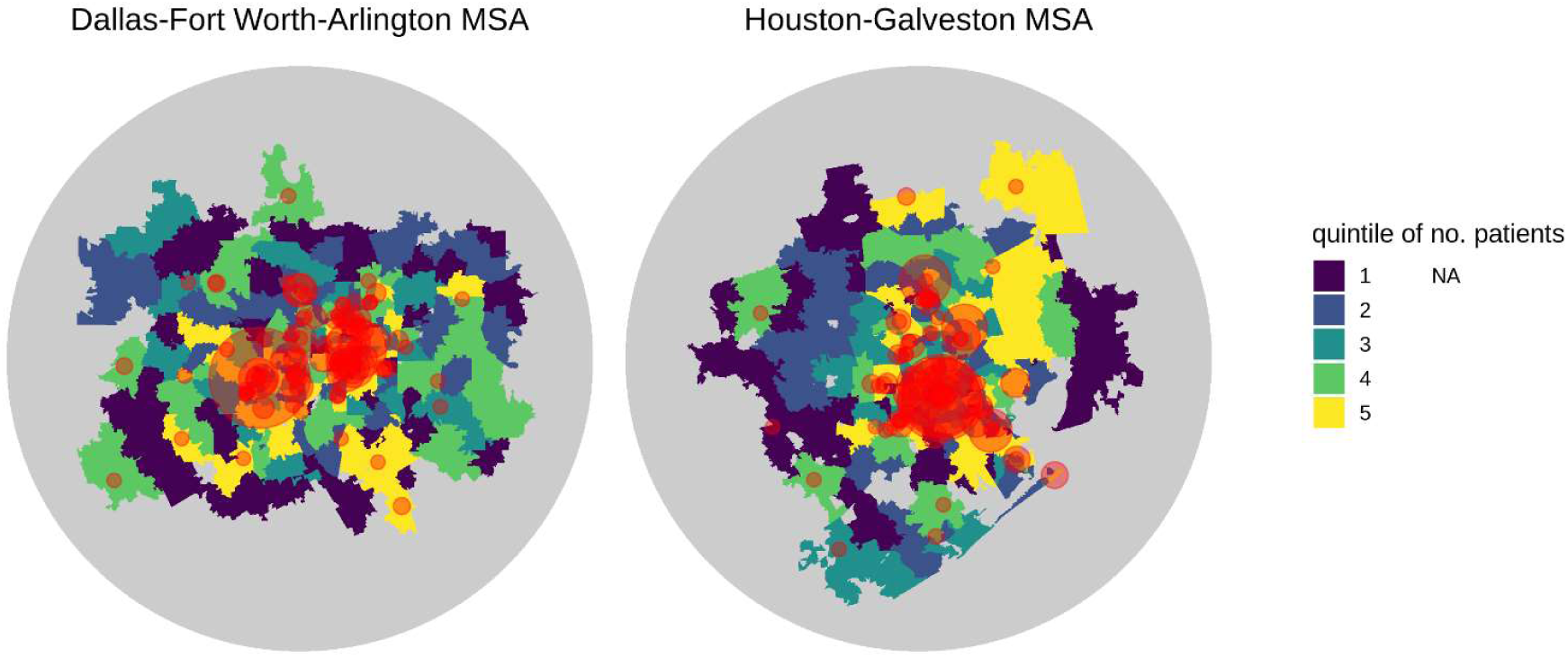
Maps of the Dallas-Fort Worth-Arlington (left) and the Houston-Galveston (right) metropolitan statistical areas as examples of a sprawling urban centers. Color coding shows quintile of absolute number of patients with symptomatic carotid disease per zi p code. Semi-transparent red circles demonstrate location of hospitals performing CEA/CAS, with circle size reflecting procedural volume. Both areas have similar rates of early carotid revascularization. The map of the Houston-Galveston does demonstrate a top-quartile region in the northwest part of the area has relatively few hospitals performing early carotid revascularization.

## Discussion

Our analysis suggests increased access to care – via increased per capita number of total hospitals and the number of hospitals performing early CEA/CAS -- will increase the number of patients receiving early CEA/CAS for carotid stenosis with stroke or TIA. In other words, decentralizing CEA/CAS procedures, not regionalizing or concentrating them. This finding may differ from literature supporting the regionalization concept for a number of specific reasons. First, CEA and CAS for stroke or TIA are much more frequently performed than other procedures such as esophagectomies for esophageal cancer or pancreaticoduodenectomies for periampullary cancers. Whereas complex cancer resections are typically done by surgeons with narrow clinical focus, CEA and CAS are more routine and done by surgeons or interventionalist physicians across a wide variety of training backgrounds.^10^ Patients with cancers that are rare or require complex resections are often referred by an internist or oncologist to a particular provider or hospital within a region known to focus on that particular problem, while, patients with stroke or TIA are much more likely to present to the closest emergency department for evaluation.

Although other analyses have found a volume-outcome correlation for CEA (not CAS), the absolute difference in combined stroke or death in high versus low volume centers is ∼2%, much smaller than that reported for pancreatectomy or esophagectomy.^11^ Arguments for regionalization of CEA/CAS need to balance the small benefit of reduced post-operative stroke and mortality rate with the increased rate of recurrent stroke and TIA occurring between initial presentation and CEA/CAS.

Our findings are consistent with other studies that have demonstrated that an increased number of hospitals or providers delivering critical medical services improve outcomes. Specifically, local or regional competition has been associated with reduced mortality among persons newly initiated on hemodialysis, reduced hemodialysis-associated hospitalizations, and increased access to liver and kidney transplantation.^12–15^ Regionalization of coronary intervention within the VA system has been associated with underutilization of angiography in acute myocardial infarction due to limited availability in remote centers.^16^ Most studies of CEA/CAS have rightly focused on individual patient-level factors associated with stroke and TIA outcomes. This study provides important insights on how geospatial and institution-level factors impact procedures done for stroke and TIA and how providers might organize services to increase patient access to CEA/CAS.

Other interventions with similar urgency and distribution would likely demonstrate a similar benefit to patient care. Increased access to fracture fixation, laparoscopic appendectomy, and coronary angiography among others would be expected to increase utilization. Coverage gaps of surgical care often disproportionately affect rural and minority communities.^17^ Often acute surgical diseases occur after a period of chronic symptoms. This interval serves as a period where access to surgical evaluation and treatment can limit emergent presentations.^18^

Of the two parameters found to be most influential, bolstering CEA/CAS procedural volumes at non-dominant hospitals would be most beneficial means of increasing the rate of early CEA/CAS rates. This would probably be best accomplished by streamlining the process of referring symptomatic carotid patients to surgeons or interventionalists and/or removing barriers surgeons or interventionalists might encounter in performing early CEA/CAS. A study by Noronen and colleagues detailing the time intervals seen between stroke or TIA presentation and CEA in Helsinki provide many actionable suggestions, including expedited vascular imaging and surgeons (or other interventionalists) performing interventions on an “on-call” or unscheduled, added to the operating room (or interventional suite) schedule to ensure the procedure is done within a two week or other abbreviated time period.^19^

We emphasize that the regression analysis, distance calculations, and qualitative analysis of maps in our study together suggest that it is indeed the balance or distribution of procedures within a region that is influential, not the location or distance. It is for this reason we are suggesting bolstering already-existing CEA/CAS services at relatively low volume (non-dominant) hospitals would be more beneficial than an expansion increasing the total number of hospitals offering CEA/CAS, though it is possible that such an expansion could have a beneficial effect as well. We would also emphasize the need for oversight or data transparency (e.g. publishing post-procedural stroke rates) to ensure that bolstering or expanding services to increase access and decreasing recurrent stroke/TIA in the interval between initial event and CEA/CAS within a region is not offset by an increased number of post-procedural neurological events.

It is possible that patients included in this analysis were discharged and scheduled for an elective procedure within two weeks. Even with this possibility, however, the relationships we have found between the rate of early CEA/CAS, number of hospitals per capita, and the procedural Gini coefficient would persist unless the proportion of patients discharged with plans for early return varied significantly among Texas MSAs. Additionally, our analysis was limited to the data through 2013 that is publicly available. The low proportion (15.4%) of patients receiving early CEA/CAS in this study is quite similar to that reported by others during a similar study period, and we hope that the proportion has increased since 2013. Even still, an increase in the proportion of early CEA/CAS would not necessarily change the validity or the importance of the relationship found in this analysis.^19^

In summary, rates of early CEA/CAS correlate strongly with hospital density and procedural distribution within a given Texas metropolitan area. Efforts to expand the number of hospitals in metropolitan areas providing carotid revascularization may provide benefit to patients with symptomatic carotid disease.

## Data Availability

All data produced in the present study are available upon reasonable request to the authors.

## Abbreviations

CAS: carotid artery stent
CEA: carotid endarterectomy
IQR: interquartile range
MSA: metropolitan statistical area

